# From Prompt to Patient: Cost-Effective Simulation Tool for Medical Education in Low-Resource Settings – Evidence from Mozambique

**DOI:** 10.64898/2026.08.13.26360321

**Authors:** Pinto Francisco Impito

**Affiliations:** Department of Arts and Humanities, Licungo University, Quelimane, Mozambique

## Abstract

Medical education in resource-limited settings faces significant challenges in providing diverse clinical exposure and fostering essential skills such as clinical reasoning, communication, and empathy. Due to the inability to afford immersive technologies such as virtual reality (VR) and Augmented Reality (AR), constrained by financial, infrastructural, and structural barriers, interactive simulation videos (ISV) constitute an innovative, cost-effective educational tool that can bridge the gap between theoretical knowledge and practical clinical experience, while enhancing student engagement and learning outcomes. This study aimed to assess the educational value and user experience of ISV as a supplementary tool in teaching medical semiology among medical students in Mozambique. A quantitative, descriptive, cross-sectional study was conducted among 4th-year medical students at Alberto Chipande University. Descriptive and inferential statistical analyses were performed, including a one-sample t-test to compare responses against a value. A total of 93 students participated in the study. The ISV was highly rated for realism (77.4%), relevance to training (74%), and usefulness of feedback (78.5%). Most students reported increased confidence in patient care (81.7) and found the digital patient credible and engaging (74.2). Overall mean scores across all domains were significantly higher than the neutral benchmark (p<0.05), indicating a positive perception of the tool. Students also expressed a strong willingness to recommend its integration into medical curricula. In conclusion, ISV represents a valuable and feasible pedagogical approach in medical education, particularly in low-resource settings. They enhance clinical reasoning, engagement, and confidence while providing scalable, standardized learning experiences. ISV holds strong potential as a complementary tool to bridge gaps in traditional medical training and improve educational equity.

## 1. Introduction

Contemporary medical education faces the challenge of balancing the acquisition of technical skills with the development of communication, empathy, and clinical reasoning abilities. Traditionally, teaching medical semiology relies on direct patient interactions, peer practice, and mannequins-fundamental strategies-but these have limitations regarding the variety of clinical cases and the ability to replicate complex real-world situations [1-4]. These limitations are even more apparent in resource-limited settings, such as many educational systems in low- and middle-income countries. [5]

Meanwhile, advances in digital technologies have led to major changes in medical education, opening up opportunities to incorporate innovative tools such as Virtual Reality (VR) [6, 7], Augmented Reality (AR) [8, 9], and Artificial Intelligence (AI) [10-12], which promote active, student-centered learning. Among these tools, digital narratives and simulation videos stand out, combining visual, auditory, and interactive elements to improve the learning experience. These methods provide exposure to various clinical scenarios, allow for repeated practice [13], [14], safe skills training [15], and help develop diagnostic reasoning in controlled settings. [16], [17]

In this context, interactive simulation videos (ISV) represent an evolution of traditional simulation methods, incorporating interactive features that place the student as the decision-maker [18]. Unlike traditional videos, ISV allows users to make clinical choices, explore different routes, and receive immediate feedback [19], thereby encouraging more reflective, personalized, and engaging learning. This method supports modern teaching models such as constructivism [20] and problem-based learning [21], which are aided by artificial intelligence [22], and emphasize active student involvement in knowledge construction.

Furthermore, integrating digital tools such as interactive simulation videos is particularly relevant within the framework of narrative medicine, which emphasizes active listening, interpreting patients’ stories, and understanding illness as a human experience [23], [24], [25]. By including clinical narratives in audiovisual and interactive formats, ISV allows students not only to identify signs and symptoms but also to grasp the patient’s emotional, social, and cultural context. This aspect helps create a more humanized education, fostering empathy and strengthening the doctor-patient relationship from the early stages of training.

On the other hand, the growing familiarity among students with digital environments, typical of younger generations [26], [27] creates unique opportunities to implement innovative teaching methods in medical education [28]. The use of interactive technologies aligns with students’ expectations while promoting autonomy, critical thinking, and self-directed learning [15]. In this way, interactive simulation videos serve not only as technological tools but also as pedagogical strategies that meet modern needs in health education [29],[15], [30], helping to bridge the gap between theory and practice and to prepare future professionals for an increasingly digital and complex clinical setting.

Although innovative digital solutions hold promise, their implementation in medical training still faces major obstacles, especially in settings with technological gaps and varying levels of digital literacy. Therefore, it is crucial to evaluate not only the pedagogical effectiveness of these tools but also students’ perceptions and the influence of these tools on clinical readiness.

This study is grounded in this framework, developed at the medical faculties of the Catholic University of Mozambique and Alberto Chipande University. The research aims to assess the educational value and interactive experience of ISV as a supplementary tool in teaching medical semiology. The goal is to enhance understanding of the role of digital technologies in medical education, especially in resource-limited settings, and to explore the potential of ISV as an innovative approach for developing clinical reasoning and decision-making skills.

## 2. Methods and material

### 2.1 Study design

This study adopted a quantitative, descriptive, and cross-sectional design to assess medical students’ perceptions of using interactive simulation videos (ISV) as a supplementary tool in medical semiology education. The quantitative approach was prioritized to enable systematic analysis of variables related to usability, perceived usefulness, and the educational impact of the ISV by collecting structured data suitable for statistical analysis. This design provided a comprehensive view of participants’ opinions at a specific point in time, identifying relevant trends and response patterns to evaluate the tool in an academic context.

### 2.2 Questionnaire design

The data collection instrument consisted of a structured questionnaire developed specifically for this study, informed by the existing literature on the evaluation of educational technologies and simulation tools in medical education. The questionnaire was designed to assess multiple aspects of the user experience, including usability, content clarity, pedagogical relevance, student engagement, and the contribution to the development of clinical reasoning. Most questions used Likert scales to quantify participants’ levels of agreement with various statements. Additionally, some questions on sociodemographic and academic characteristics were included to aid interpretation of the results.

A pilot test was conducted to assess the questionnaire’s clarity, comprehensibility, and suitability. The pilot was performed with 3rd- and 4th-year students from the medical schools of the Catholic University of Mozambique (UCM) and Alberto Chipande University (UNIAC). The participation of these students helped identify potential difficulties and ambiguities in the questionnaire. Based on the feedback received, necessary revisions were made before moving to the data collection phase.

### 2.4 Recruitment and data collection

Participants were recruited from medical students at the participating institutions via convenience sampling. The invitation to participate was distributed through informal communication channels within the classes, namely, WhatsApp groups used by the medical students, enabling quick and effective dissemination of the study. After the invitation, participants had access to the interactive simulation video and were subsequently asked to complete the online questionnaire. Data collection was conducted digitally, ensuring easy access and flexible participation. Participation was voluntary, and students were informed in advance of the study’s objectives and the confidentiality and anonymity of their responses. Only fully completed questionnaires were included in the analysis.

### 2.5 Ethical approval

The study was conducted in accordance with applicable international ethical principles for research involving human subjects. The research received approval from the National Bioethics Committee for Health in Maputo on November 17, 2024, under registration number 54/CNBS/2024, confirming its compliance with the ethical requirements established for this type of investigation. All participants were properly informed about the nature, objectives, and procedures of the study, and informed consent was obtained prior to their participation. Anonymity and confidentiality of the collected data were ensured, and the data were used exclusively for scientific purposes. Participation was entirely voluntary, with the option to withdraw at any time without penalty to the participants.

## 3. The simulation tool design

This section encompasses four intervention stages considered crucial to the study’s effective conduct. There are four stages: stage 1, which corresponds to the brainstorming moment; stage 2, video production, which includes the process of its creation; stage 3, interactive tool creation, which includes branching and interactivity; and stage 4, tool usability, dedicated to assessing the IVS usability prior to its implementation.

### Stage 1: *Brainstorming*

This initial stage involved defining and presenting a proposal for an interactive simulation video for medical students. It included a panel comprising the principal investigator, who serves as an audiovisual and interactive content producer, and two medical faculty lecturers from Alberto Chipande University, forming a multidisciplinary team. This stage lasted one month, from March to April 2025, covering planning, instructional design, and the development of pedagogical specifications that guided the VSI’s development.

The meetings held at this stage included developing the patient clinical story via an AI model (DeepSeek), based on suggestions collected during the first meeting, as well as a simple computer-based prototype to illustrate the VSI interface (Figures 1 and 2).

**Figure 1.**
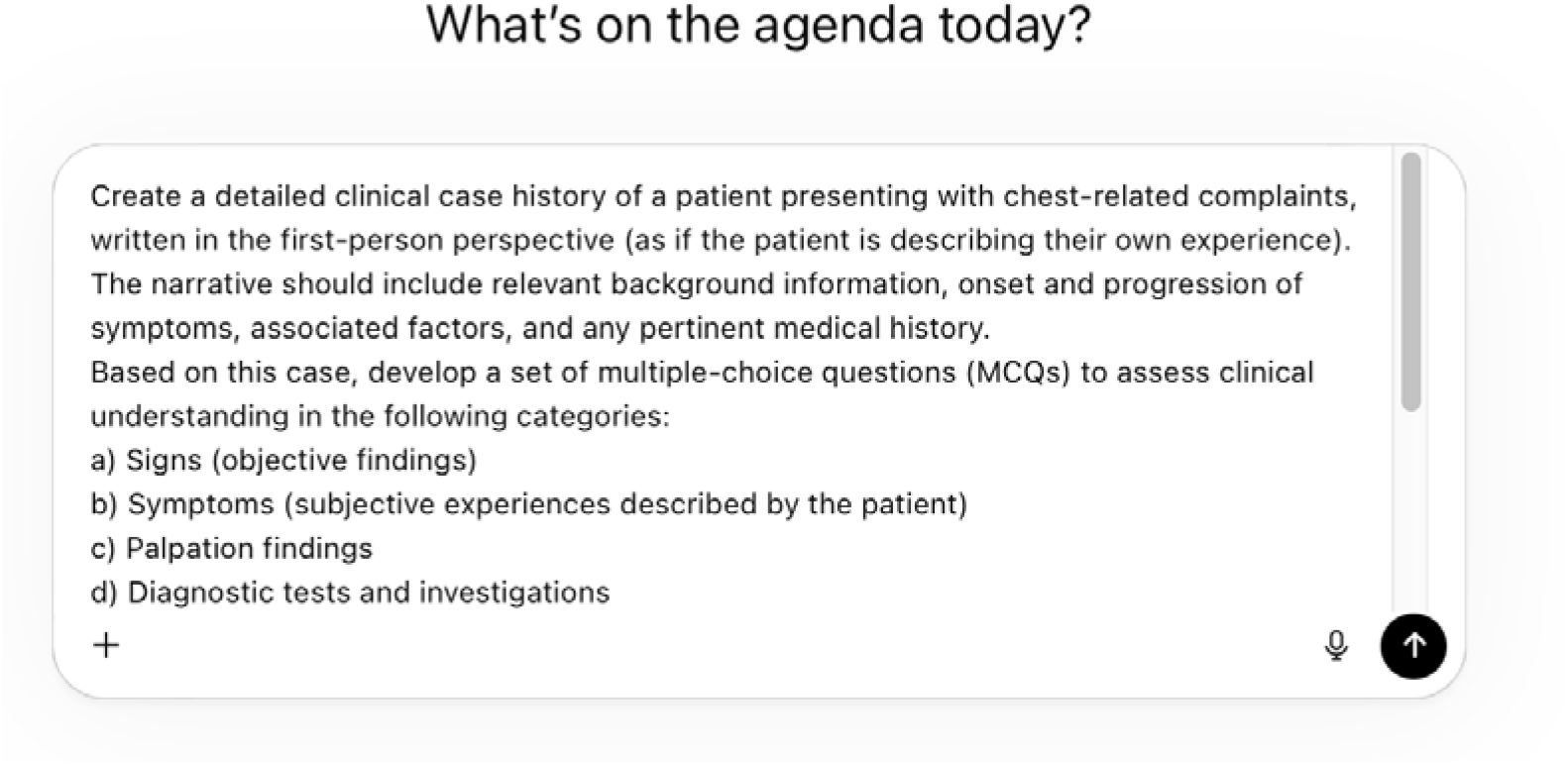
Prompts used to generate the patient’s clinical story **Source:** ChatGPT

**Figure 2.**
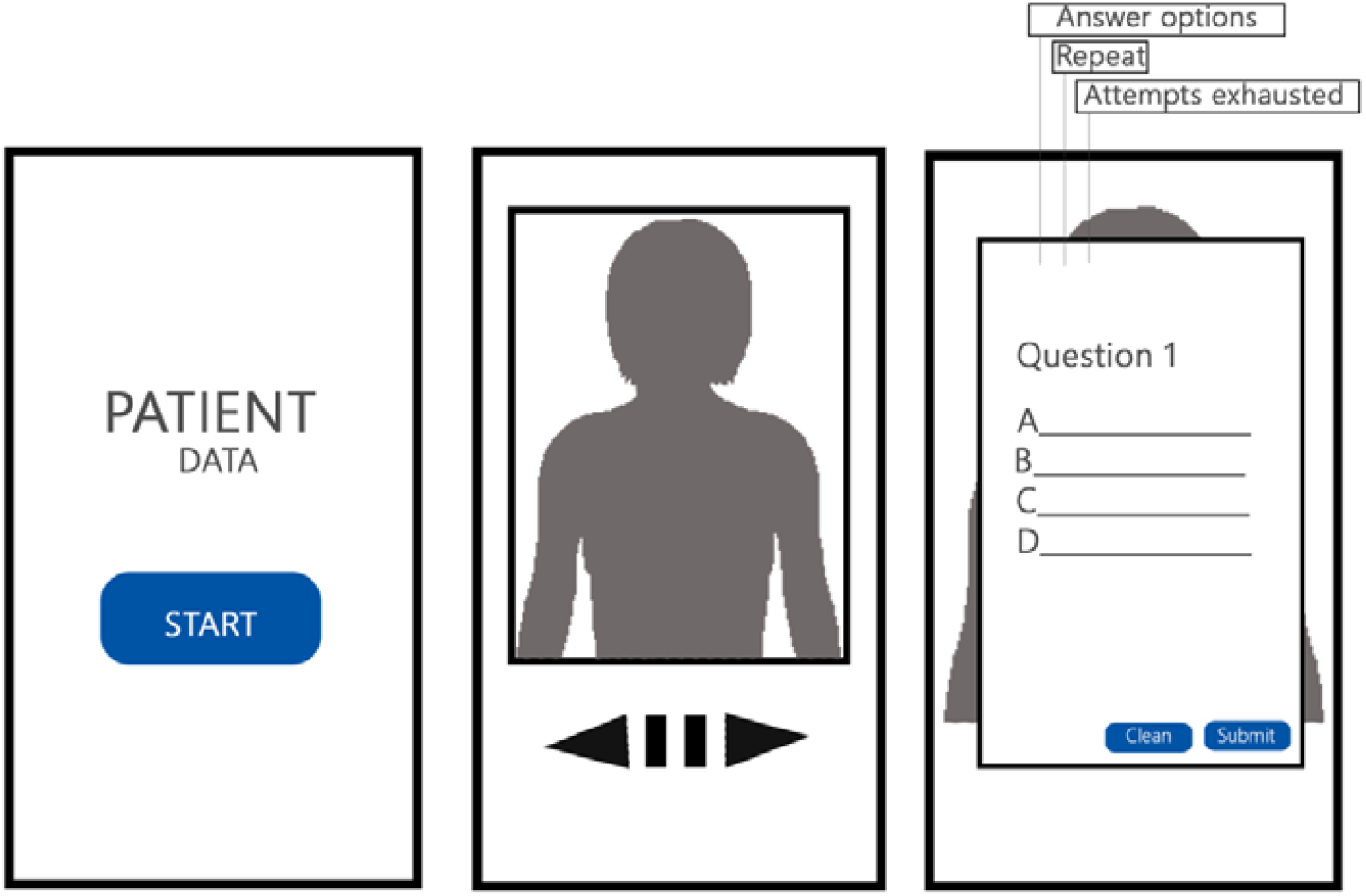
Handmade prototype extract for illustration purposes

**Figure 3.**
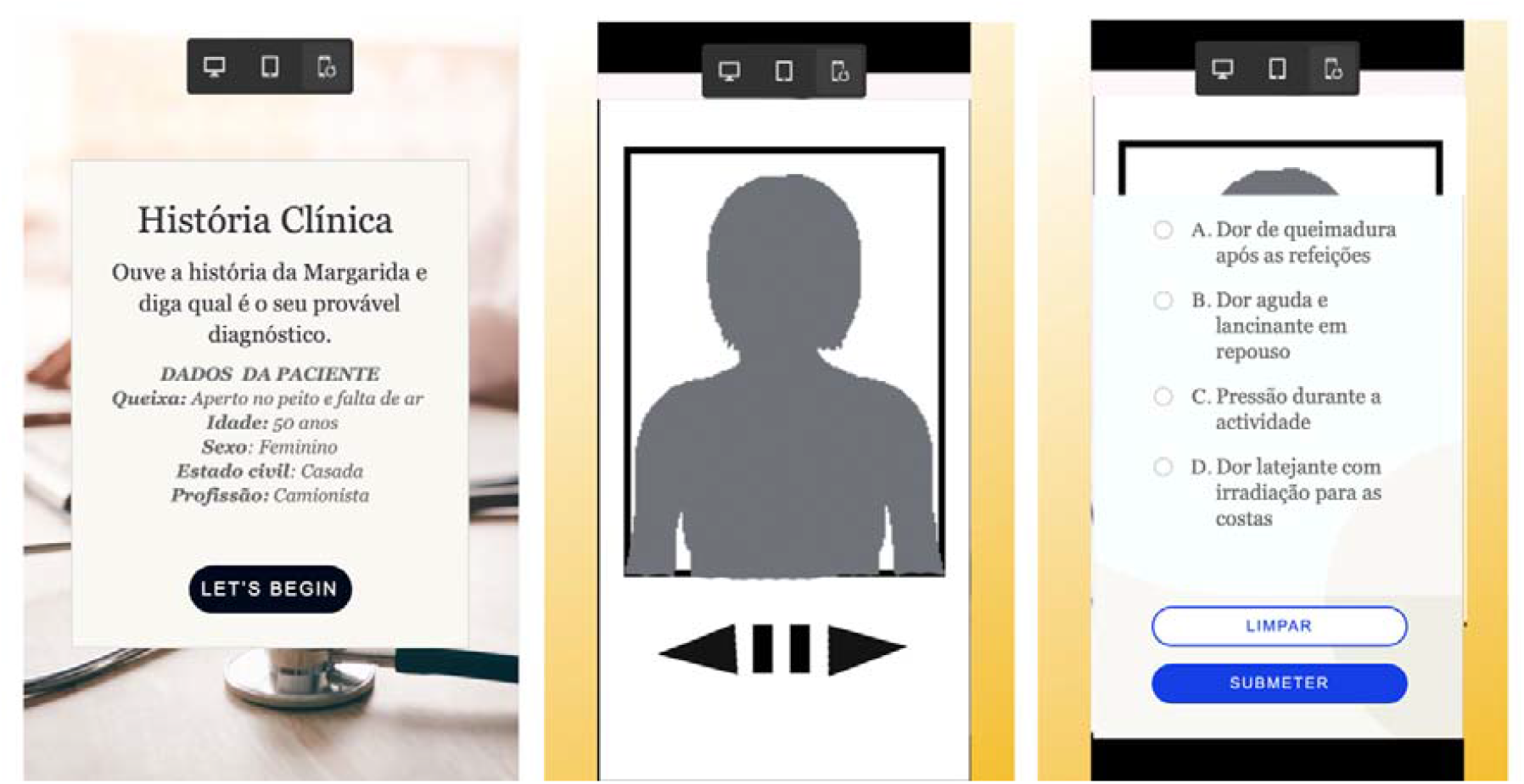
Creation of Interactive Simulation Video in Adobe Captivate

The final clinical history, AI-generated (supplement 1), including the multiple-choice questions, was also presented in this stage and approved accordingly. The use of artificial intelligence to develop practical exercises in medicine is common, especially to improve understanding of diagnosis, treatment, and disease prevention. [31]

The developed prototype was presented to lecturers to familiarize them with the interface and assess its pedagogical suitability. During its development, essential user experience principles such as ‘mapping’ and ‘affordance’ were considered, according to Don Norman. [32]

### Stage 2: *Video production*

As a result of the thorough planning carried out during phase 1, this new stage involved the researcher (cameraman and director) and the actress portraying the patient, whom Bates considers capable of playing this role convincingly [33], alongside a boom operator responsible for positioning the microphone close to the actress, out of frame, and a gaffer in charge of controlling the lighting, both in terms of intensity and direction (Table 1).

**Table 1.**
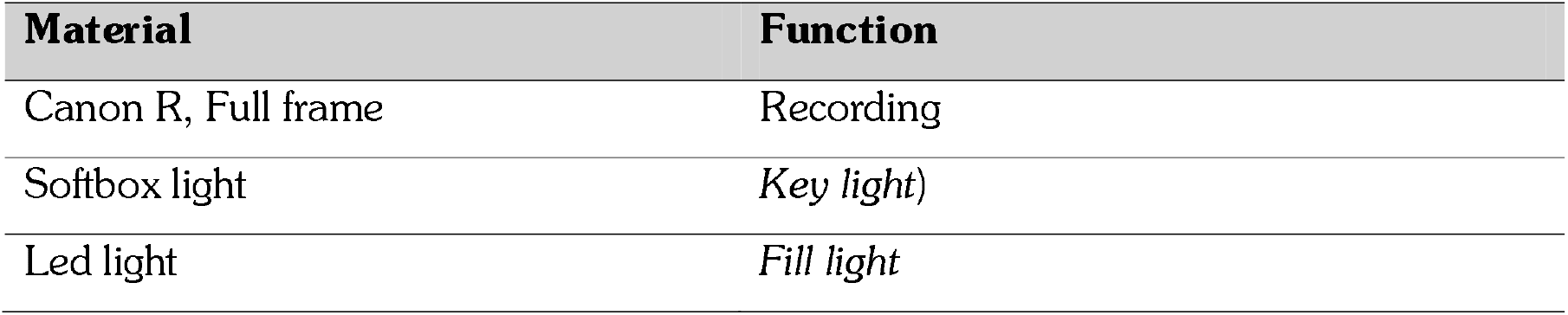
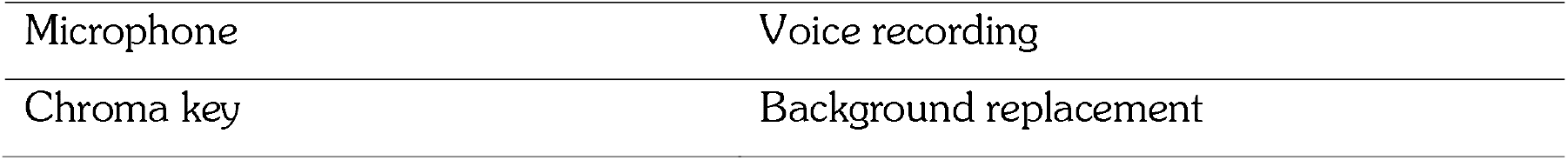
Description of the materials used.

| Material | Function |
| --- | --- |
| Canon R, Full frame | Recording |
| Softbox light | Key light) |
| Led light | Fill light |
| Microphone | Voice recording |
| Chroma key | Background replacement |

According to Bates, an actor chosen to portray content related to health education may be someone who probably has not experienced what they are representing but can recreate it convincingly [33]. The production was divided into four main steps: (1) rehearsal, focused on expression, intonation, and naturalness in acting; (2) recording, which captured the necessary images and sounds; (3) editing, which included selecting, organizing, and processing the images to ensure coherence, continuity, and pedagogical effectiveness; and (4) tool design, which involved integrating the video (clinical history) into a platform that made it interactive.

### Stage 3: *Interactive tool creation*

This final phase involved transforming the edited video into an interactive tool that provides immediate feedback exercises and supports continuous user engagement. Interactivity was implemented in Adobe Captivate through a dynamic process that enabled logical branching across different levels of interaction. The branches refer to the programmed links between interface elements, such as the ‘start,’ ‘stop,’ ‘clear,’ and ‘submit’ buttons, as well as the response options to the questions presented during the simulation.

This interactive design offers an intuitive pathway with simple tasks that promote speed and efficiency, while providing personalized experiences and reinforcing active learning through the simulation of clinical decision-making, which is essential for developing skills in medical education. After the tool (the VSI) was created, it was presented at a forum for medical educators, who evaluated it positively. This preview and the functional tests conducted with educators also aimed to confirm that the buttons (start, stop, clear, and submit) and the transitions between content levels functioned correctly. This verification phase was crucial to ensure that navigation was smooth, the branches were logically consistent, and the interface responded correctly to user choices.

### Stage 4: *Tool usability*

The usability assessment consisted of an intervention stage conducted prior to the main participants’ use of the ISV (quantitative study). It involved a small group of 3rd and 4th-year students from the Medical Schools of Alberto Chipande University and the Catholic University of Mozambique. Its objective was to evaluate the formative usability of the interactive simulation video (ISV), focusing on user experience, interface clarity, levels of interaction, and the tool’s overall design.

This stage was divided into two substages, which are (1) initial testing and (2) iteration. The first, termed the initial stage (involving 3rd-year students), was conducted to test the tool, identify errors, and implement corrections. The second, the iteration stage (involving 4th-year students), aimed to confirm that errors identified in the first stage had been successfully resolved.

The engagement sessions took place in classrooms with students from their respective universities, who interacted with the tool in groups of three or four, using a local computer to access Adobe Captivate via a generated link. This was followed by a semi-structured interview also conducted in groups. Among the main observations from the initial testing phase, students first valued the tool, as indicated by the word cloud in Figure 4.

**Figure 4.**
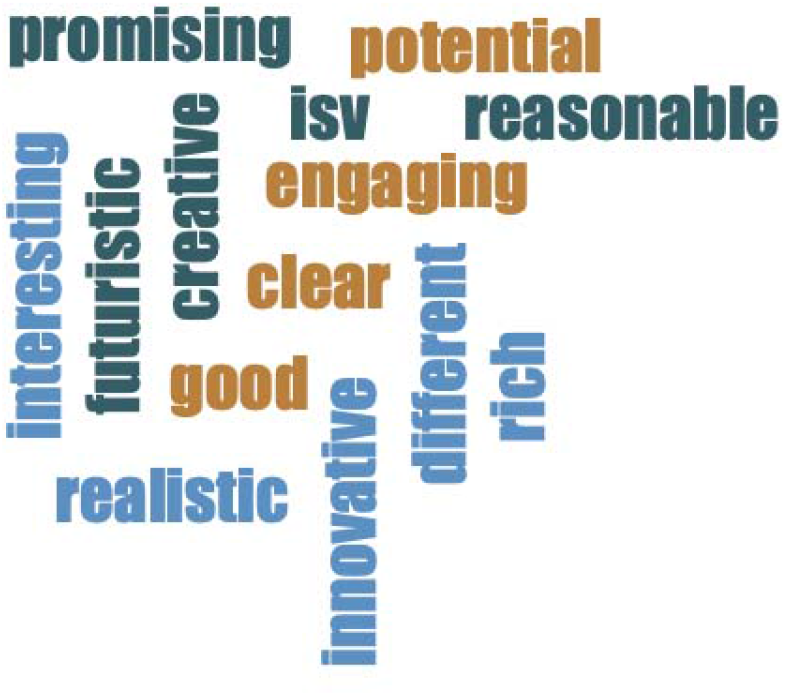
Word cloud illustrating participants’ perceptions of the tool following usability testing

After that, the following observations stood out: (1) the emergence of questions before the presentation of the necessary information to answer them, and (2) the high number of questions throughout the interaction, which prolonged the duration of the simulation. On the other hand, observations regarding the behavior of the ‘submit’ button (color change), the absence of photos illustrating the patient’s skin, and the recommendation that the patient be more spontaneous rather than rigidly follow a script were noted. The previous two identified errors were corrected and confirmed during the iteration phase. The observations regarding the submission button and the overall performance were not considered obstacles that could preclude the tool’s eligibility to proceed to assessment with the larger student sample.

Based on feedback collected during the usability assessment, improvements were implemented to the content structure, interface, and video progression logic. During the iteration phase, a significant increase was observed in ease of use, understanding of clinical scenarios, and participant engagement, reflecting a better adaptation of the tool to users’ needs. Thus, the iterative process helped optimize the usability of the VSI, reinforcing its potential as an effective tool for testing with the broader public.

Finally, the ISV was implemented in the quantitative study using the same access approach used in the usability test. Students accessed the tool on their mobile phones via a link from Adobe Captivate in a classroom that accommodated 20 students per day, prior to completing the questionnaire.

## 4. Data management and statistical analysis

The data collected through the online questionnaire were exported to a digital database and subsequently organized and processed for analysis. Initially, a check was performed to verify the data’s consistency and integrity, with incomplete or inconsistent responses excluded. Statistical analysis was conducted using SPSS version 30 to handle quantitative data, enabling descriptive and inferential statistics. Categorical variables were described using absolute and relative frequencies, while continuous variables were presented as means and standard deviations, or medians and interquartile ranges, depending on the data distribution.

To assess the internal consistency of the questionnaire, Cronbach’s alpha was calculated, yielding 0.76, indicating acceptable reliability. Descriptive statistical analysis was performed using frequency, percentages, means, and standard deviations to summarize participants’ responses. For inferential analysis, a one-sample t-test was used to determine whether the mean score for each item differed significantly from the neutral value on the Likert scale (reference value = 3). The level of statistical significance was set at 5% (p<0.05). Additionally, 95% confidence intervals were estimated for each mean to assess the precision of the results. All findings were presented in tables to facilitate the interpretation and communication of the results.

## 5. Results

### 5.1 Socio-demographic characteristics

In total, 93 students participated in the questionnaire, out of the 95 who interacted with the Interactive Simulation Video (ISV). The questionnaire was answered exclusively by the target audience for this study, namely 4th-year students in the Faculty of Medicine at Alberto Chipande University. Among these students, 65.6% were female, and 34.4% were male. Regarding age groups, the distribution was relatively balanced, with 33.3% aged 18-24, 40.9% aged 25-35, and 25.8% aged 36 or older.

### 5.2 Descriptive analysis

#### 5.2.1 Educational value of ISV

Regarding this variable, the results were significantly positive, with more than half of the participants (N=72, 77.4%) completely agreeing that the simulation reproduced realistic clinical situations, and (N=69, 74.2%) that it was relevant for medical training. The usefulness of corrective ‘feedback’ after incorrect responses was highly valued (N=73, 78.3%). The structured action-feedback-reflection sequence received a lower rating compared to other Likert items (N=55, 59.1%) (Table 2).

**Table 2.**
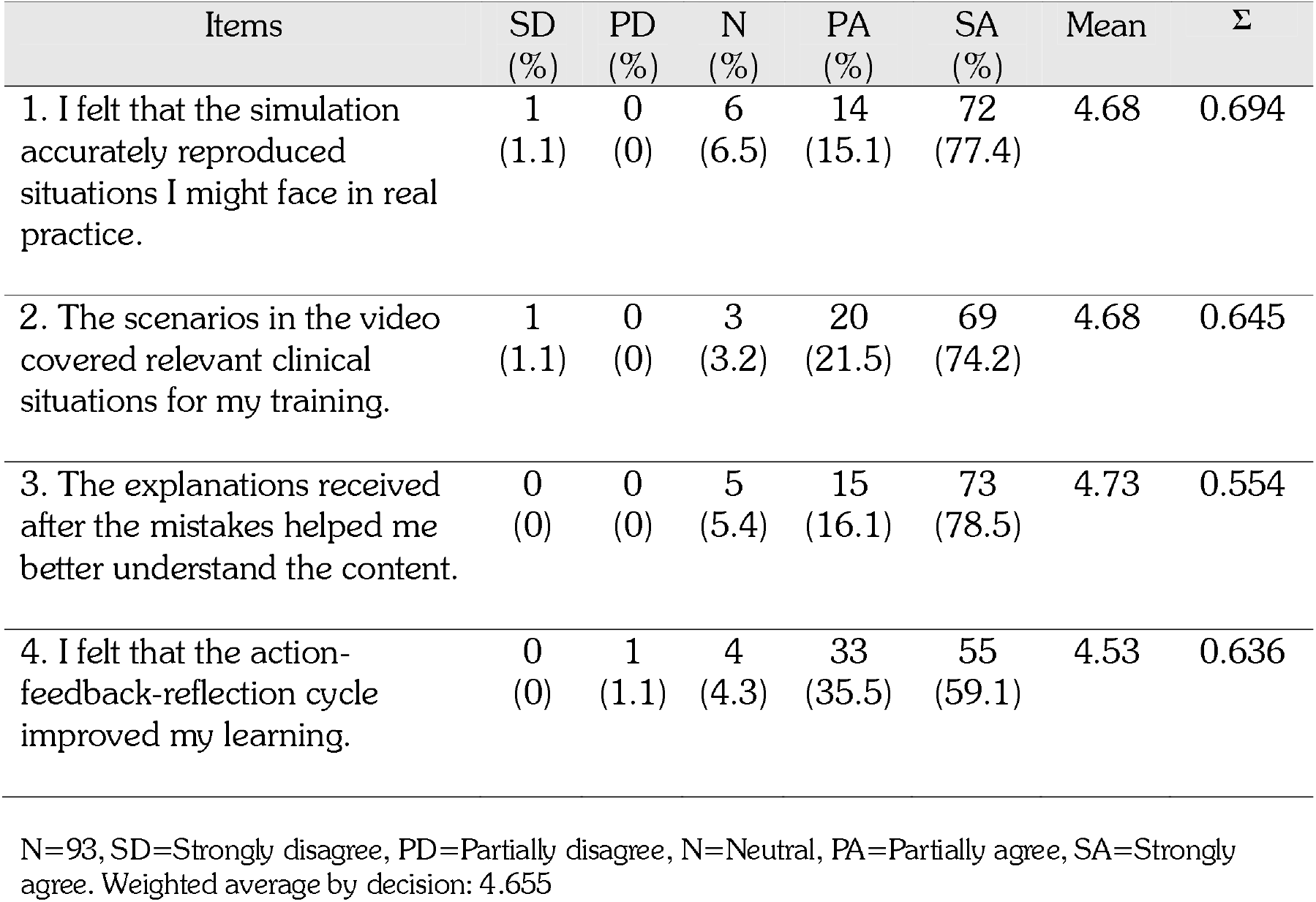
Educational value of ISV.

Regarding this variable, the analysis conducted revealed that most participants evaluated the tool positively, indicating that the simulations strengthened their confidence in clinical care (N=76, 81.7%); the digital patient and the naturalness of interaction with the content were considered credible (N=69, 74.2%); and the level of recommendation of the tool for medical courses was quite high (N=83, 89.2%). Conversely, the Likert item regarding the possibility that the ISV was considered more realistic received a low rating (N=58, 62.4%), as did the level of participants’ emotional engagement throughout the entire experience (N=59, 63.4%) (Table 3).

**Table 3.** Appreciation of interactive experience with VSI.

| Items | SD<br>(%) | PD<br>(%) | N<br>(%) | PA<br>(%) | SA<br>(%) | Mean | Σ |
| --- | --- | --- | --- | --- | --- | --- | --- |
| 1. These simulations are important to increase confidence in patient care. | 0<br>(0) | 0<br>(0) | 0<br>(0) | 17<br>(18.3) | 76<br>(81.7) | 4.82 | 0.389 |
| 2. The simulation was so realistic that it made me forget it was simulated. | 2<br>(2.2) | 0<br>(0) | 6<br>(6.5) | 27<br>(29.0) | 58<br>(62.4) | 4.49 | 0.802 |
| 3. The simulation kept me emotionally engaged throughout the entire experience. | 2<br>(2.2) | 0<br>(0) | 1<br>(1.1) | 31<br>(33.3) | 59<br>(63.4) | 4.56 | 0.729 |
| 4. A paciente digital e as interações utilizadas foram credíveis e contribuíram para o meu envolvimento. | 0<br>(0) | 0<br>(0) | 1<br>(1.1) | 23<br>(24.7) | 69<br>(74.2) | 4.73 | 0.469 |
| 4. The digital patient and the interactions used were credible and contributed to my engagement. | 1<br>(1.1) | 0<br>(0) | 1<br>(1.1) | 8<br>(8.6) | 83<br>(89.2) | 4.85 | 0.531 |

N=93, SD=Strongly disagree, PD=Partially disagree, N=Neutral, PA=Partially agree, SA=Strongly agree. Weighted average by decision: 4.655

### 5.3 Complementary Inferential Analysis

Beyond descriptive statistics, an inferential analysis was conducted to assess whether the means across the evaluated dimensions significantly differed from the neutral value on the Likert scale (3). For this, a one-sample t-test was applied, considering a significance level of 5% (p < 0.05). The results showed that all observed means, across both the educational value and interactive experience dimensions, were significantly higher than the neutral scale value. These findings indicate that students had a statistically positive perception of the interactive simulation video, reinforcing its potential as an effective pedagogical tool in teaching medical semiology (Table 4).

**Table 4.** Inferential analysis of students’ perceptions of the interactive Simulation Video (One-Sample t-test)

| Item | Mean | SD | t | p | 95% CI |
| --- | --- | --- | --- | --- | --- |
| Educational value |  |  |  |  |  |
| Realism of the simulation | 4.68 | 0.694 | 27.1 | <0.001 | 4.54 – 4.82 |
| Relevance of the simulation | 4.68 | 0.645 | 28.1 | <0.001 | 4.55 – 4.81 |
| Feedback after errors | 4.73 | 0.554 | 30.7 | <0.001 | 4.62 – 4.84 |
| Action-feedback-reflection cycle | 4.53 | 0.636 | 22.7 | <0.001 | 4.40 – 4.66 |
| **Interactive Experience** |  |  |  |  |  |
| Confidence in patient care | 4.61 | 0.662 | 24.5 | <0.001 | 4.47 – 4.75 |
| Perceived realism | 4.67 | 0.613 | 26.3 | <0.001 | 4.54 – 4.80 |
| Emotional involvement | 4.64 | 0.598 | 25.7 | <0.001 | 4.52 – 4.76 |
| Credibility of patient/interaction | 4.59 | 0.634 | 24.1 | <0.001 | 4.46 – 4.72 |
| Recommendation of the tool | 4.72 | 0.552 | 31.0 | <0.001 | 4.61 – 4.83 |

Additionally, 95% confidence intervals were estimated for the means, allowing assessment of the precision of the estimates obtained. The calculated intervals did not include the neutral scale value, corroborating the statistical test results and strengthening the robustness of the conclusions.

## 6. Discussion

The results of this study highlight a generally positive perception among medical students regarding the use of interactive simulation videos (ISV) as a supplementary tool in medical education. Consistently, the participants recognized its pedagogical value, particularly in supporting the development of clinical reasoning, enhancing engagement in the learning process, and providing opportunities for interaction with simulated clinical scenarios.

These findings support previous evidence emphasizing the role of interactive technologies in promoting active, student-centered learning aligned with Fosnot’s constructivist principles and Ausubel’s concept of meaningful learning, which holds that learning occurs through the active reconstruction of knowledge, mediated by meaningful experiences [34, 35], enhancing confidence [1], and bringing students closer to real clinical situations. [27]

The analyses reported increases in students’ confidence and in their emotional and technical readiness to treat real patients after using the VSI. Additionally, there was a perception that the VSI could contribute to students’ clinical preparation and reduce anxiety and fear of making mistakes when working with patients by providing problem-based learning (PBL) [36, 37]. By sharing their story in the first person, the digital patient not only conveys important clinical data but also creates an emotional connection, promoting person-centered learning. This approach aligns with the principles of Narrative Medicine, as emphasized by authors such as Charon [23], Kleinman 12], and Shapiro [25], who emphasize the importance of active listening, empathy, and the human aspect in clinical practice.

The question aimed to understand how the use of the VSI would influence students’ readiness for future clinical practice in a context of increasing integration of digital technologies, revealing that students’ contact with the VSI fostered a deeper understanding of the role of digital solutions in decision-making, aligning with three key learning factors: observation, reflexion, and action, as suggested by Bandura [38], Kolb [39], and Vygotsky [40]. Therefore, the importance of incorporating this technology into medical education in a continuous, up-to-date manner is emphasized not just as an additional resource but as a fundamental part of the teaching and learning processes. [41], [42], [43]

The expressed interest in integrating the VSI into the medical school curriculum, as well as the possibility of repeating exercises and reviewing clinical signs, aligns with studies highlighting the importance of hybrid curricula in developing countries, where digital simulations can compensate for limited physical and human resources [44],[45],[46],[47], fostering critical thinking, encourage autonomy, and make students active participants in the learning process. [15], [13], [14].

In the specific context of Mozambique, where structural challenges and resource limitations in clinical training are prevalent, the ISV emerges as an innovative solution that can complement traditional education. The possibility of remote access, combined with repeated clinical scenarios and standardized learning experiences, can help reduce inequalities in access to clinical training and improve students’ preparedness for professional practice.

### 6.1 Limitations and future directions

Despite the significant contributions of this study, some limitations should be considered when interpreting the results. First, the use of convenience sampling may introduce selection bias, limiting the generalizability of the findings to other medical student populations. Second, the cross-sectional design of the study does not allow for assessment of the impact of the VSI over time, particularly regarding knowledge retention or the transfer of skills to real clinical practice. Additionally, reliance on self-reported data may be subject to perception and social desirability biases and may not necessarily reflect objective changes in students’ performance.

Future research should consider adopting more robust methodological designs, including longitudinal studies and controlled trials, to evaluate the impact of the VSI on objective indicators of clinical performance. Furthermore, expanding the VSI to other areas of medical training and adapting it to different institutional and geographical contexts is recommended. Incorporating advanced features, such as adaptive feedback or artificial intelligence, could enhance personalized learning and increase the tool’s pedagogical effectiveness. Finally, it will be important to assess the feasibility of integrating the VSI into formal curricula to ensure its sustainability and long-term impact.

### 6.2 Conclusion

This study highlights that interactive simulation videos are an innovative and relevant pedagogical tool in medical semiology education, widely accepted by students and perceived as facilitators of clinical reasoning, decision-making, and engagement in the learning process. Its interactive and multimodal nature enables the creation of more dynamic and contextualized learning environments, bringing students closer to realistic clinical scenarios and promoting more effective integration of theory and practice. In a context such as medical education in Mozambique, marked by structural challenges and limited access to immersive technologies, incorporating cost-effective digital solutions, such as ISV, is particularly relevant.

This approach not only contributes to democratizing access to quality learning experiences but also enables standardization of content and repetition of clinical scenarios, key factors for skill consolidation. Thus, VSI presents itself as a complementary strategy with the potential to enhance equity and effectiveness in medical education in resource-limited settings. Beyond its immediate value as a teaching support tool, this tool is part of a broader trend of digital transformation in health education that favors student-centered methodologies, active learning, and interactive technologies. Its gradual integration into medical curricula could help train professionals better prepared to face the increasing complexity of contemporary health systems, characterized by dynamic clinical environments, technological demands, and the need for informed decision-making.

Despite the identified limitations, the results of this study reinforce the potential of interactive simulation videos as a robust and adaptable complementary pedagogical approach. Continued research in this area, combined with its gradual and sustained implementation, could solidify the role of these tools in medical training, contributing to improved teaching quality and, ultimately, better healthcare services.

## Data Availability

All data produced in the present study are available upon reasonable request to the authors

